# Traditional Healers’ Knowledge, Attitudes, and Perceptions About Tuberculosis and Collaboration with the Conventional Health System in the Kereyu Pastoralist Area of Ethiopia: A Cross-Sectional Study

**DOI:** 10.1101/2024.04.04.24305249

**Authors:** Bezawit Temesgen Sima, Tefera Belachew, Gunnar Bjune, Fekadu Abebe

**Author notes:** Corresponding author Postal address: Hadelandsveien 1294, Hakadal, Norway. <u>Authors’ addresses</u> Fekadu Abebe (BSc, Msc, PhD), Telphone, Tefera Belachew, (MD, MSc, DLSHTM, PhD), Telphone: 0251 47111461, Gunnar Bjune (MD, PhD).

## Abstract

**Background:** Traditional Healers (THs) hold significant roles in many developing countries, often sought for ailments like tuberculosis (TB). However, their knowledge, attitude, and practice (KAP) regarding TB, particularly in Ethiopia’s pastoralist areas, remains unexamined. This study evaluates THs’ KAP on TB and their perceptions to collaborate with conventional health systems on TB control.

**Method:** A cross-sectional survey was conducted among THs in Kereyu, Ethiopia from September 2014 to January 2015. Using a semi-structured questionnaire, 268 THs were interviewed. Health Extension workers helped identify the THs.

**Results:** Of the 268 participants, 80.6% were male. 97.4% were aware of TB (locally “dukubba soombaa”), with 80.2% associating its cause to proximity with a TB patient. Coughing for over two weeks was identified as a primary TB symptom by 87.35%. However, 66.4% displayed limited biomedical knowledge on TB. A notable 38.4% associated TB with sadness and hopelessness, while 47.8% utilized plant-based remedies for treatment. Impressively, 86.2% expressed willingness to collaborate with conventional health services for TB control.

**Conclusion:** The THs had limited biomedical knowledge and some misconceptions about TB. Despite providing traditional medicine to treat TB, their readiness to collaborate with established health systems is promising. Thus, Ethiopian TB control initiatives should consider integrating THs via targeted training and health education interventions.

## Background

Recent estimates indicate a significant global exposure to Mycobacterium tuberculosis, with over 10 million individuals developing active tuberculosis disease annually, leading to substantial morbidity and mortality ^1^. A substantial proportion of TB diagnoses occur in Africa, reflecting the disease’s impact on marginalized populations ^2^. In Ethiopia, tuberculosis remains a major public health concern and a significant cause of death^3, 4^.

The current battle against tuberculosis is guided by the World Health Organization’s “End TB Strategy,” which sets ambitious targets to achieve a 90% reduction in TB deaths and an 80% reduction in TB incidence by 2030^1^. Despite these goals, worldwide efforts are encountering numerous obstacles, including suboptimal detection of cases, a shortage of healthcare professionals, stigma, and widespread misunderstandings surrounding TB’s transmission and treatment, issues that are especially prevalent in developing countries. These issues have been exacerbated by the COVID-19 pandemic, which has further strained healthcare systems and diverted critical attention and resources from TB control efforts^2, 4^. Traditional healers, frequently sought after in these regions, may unintentionally delay accurate TB diagnoses because of their limited understanding of the disease, thereby exacerbating the risk of dissemination and more severe TB complications^5–7^.

In Ethiopian context, a TH is typically someone believed to possess curative powers for various ailments, stemming from religious beliefs, supernatural influences, personal experience, or even hereditary factors^8^ ^9^. The nation formally acknowledged traditional medicine in 1942^8^. Prominent among them are herbalists, spiritual healers, bonesetters, birth attendants, tooth extractors, and so-called witch doctors^8, 9^.

For effective TB control, it’s proposed that national programs should focus on educating THs about TB and possibly assimilate them into mainstream healthcare via referral systems^10–12^. Previous attempts to merge THs with TB control in developing regions have highlighted the possibility of such collaboration^10, 12, 13^. However, bridging modern healthcare and traditional healing remains intricate due to misunderstandings, communication gaps, and trust issues^5, 14–16^.

Given this context, our study aims to assess the THs’ knowledge, attitude and practice regarding TB and their perception about collaboration with the conventional healthcare system on TB control efforts in the Kereyu pastoralist area. The data generated could be used to plan advocacy, communication, and social mobilization to help facilitate the collaboration of conventional health system and THs on TB prevention and care in a pastoralist area.

### Specific objectives

1. To assess knowledge of THs regarding the transmission, diagnosis, and treatment of tuberculosis in the Kereyu pastoralist area.
2. To evaluate the attitudes of THs towards tuberculosis and individuals affected by the disease in the study area.
3. To explore the perceptions of THs about their role in the management of tuberculosis and their willingness to collaborate with the conventional healthcare system.
4. To provide recommendations for integrating traditional healers into the broader tuberculosis control strategy in the Kereyu pastoralist area

## Methods

### Study Location and Population

#### Study Location

This research took place in the Fentalle (Kereyu) Wereda in the East Shoa Zone of Oromia, part of Ethiopia’s northern Rift Valley region. Spanning an area of 1,170 km^2^, this district is inhabited by approximately 76,367 individuals and lies 200 km east of Addis Ababa, with Metehara as its administrative hub. The predominant ethnicity is the Oromo who lead pastoral way of lives. This Wereda is subdivided into 18 rural kebeles (the smallest administrative units), with 15 specifically being pastoralist. The study focused on kebeles like Kobo, Benti, Dhabiti, D/Hadu from the pastoralist sectors and Alge and G/Dima from the settled areas.

The pastoralist communities are broadly categorized into two: nomadic pastoralists, who exclusively rear livestock and are without a fixed dwelling, and agro-pastoralists who blend farming with migratory livestock rearing. These pastoralists tend to move seasonally, often relocating to different sub-districts during drier times^17^.

In the district, TB, locally known as “Dhukubba Soombaa” (translating to “lung disease”), ranks as a primary health concern as per district TB report (unpublished), but no detailed TB prevalence study exists yet.

#### Definition of “Traditional Healers”

Herbalists, locally called Qoorichota Aaadaa, are the most prevalent group. They utilize wild plants, including roots and leaves, to address common health issues such as intestinal parasites and snakebites. Bonesetters, known locally as Dhidhibaa or Dhidibtuu, specialize in setting fractured bones, extracting teeth, and performing uvulectomies. Additionally, there are traditional midwives, referred to as Deesisttu Dadaa, who provide support to women during childbirth. These midwives are highly esteemed as community leaders, and their opinions are deeply respected, as gathered from discussions with Health Extension Workers.

### Ethiopian Health Tier System and Collaboration with THs

In Ethiopia, the health care system is tiered into three distinct levels. The Primary Health Care Unit forms the base, comprising health posts, health centers, and primary hospitals, designed to serve around 25,000 people. Health posts, staffed by Health Extension Workers and varying other health professionals, offer essential and preventive health services, referring more complex cases to health centers. Health centers provide broader preventive and curative care and training for HEWs, while primary hospitals deliver comprehensive inpatient, ambulatory, and emergency surgical services for approximately 100,000 people. The second tier includes general hospitals, which offer similar services to primary hospitals, act as referral centers, and provide training for medical staff, catering to about 1 million people. At the apex, the third tier includes specialized hospitals offering advanced care for around 5 million people and serving as referral centers for general hospitals^18^.

In pastoralist zones, including Kereyu, the limited accessibility to healthcare services coupled with a seasonal migration and scant understanding of TB^6^ often leads to the populace seeking the assistance of traditional healers for ailments, including TB^19, 20^. Despite the community’s limited biomedical knowledge grasp on TB, the district’s healthcare professionals, especially the HEWs, express a willingness for collaboration with traditional healers^12^. Therefore, assessing the THs’ understanding of TB, their perspective on it, and their existing TB management practices is vital for the district’s TB control program and similar environments. This assessment will facilitate the design of tailored training programs for THs on TB. In turn, this could bolster TB prevention and care by fostering evidence-based partnership between THs and the standard health system, especially in the identification and detection of presumptive TB cases.

### Study Design

Between September 2014 and January 2015, we carried out a cross-sectional survey in the community. This study focused on Traditional Healers (THs) living in a district made up of 15 pastoralist and three sedentary villages. Notably, those sedentary villages house pastoralists who transitioned to a sedentary lifestyle, coexisting with other community members. Every TH in the district, known to offer traditional healing during our study timeline, was considered. To identify these THs in each village, we collaborated with local kebele leaders and Health Extension Workers (HEWs) from the district. These HEWs, responsible for the Health Extension Program’s household implementations, have intricate insights into the village inhabitants.

### Sampling Method

Based on a single population proportion formula^21^, we determined an approximate minimum sample size of 192. This calculation considered a 95% confidence level, a 5% margin of error, and a 74.3% expected awareness level about persistent cough being a primary symptom of pulmonary TB. This expectation was derived from a study assessing pastoralists’ perception of TB in Ethiopia’s Afar region^22^. While the district’s total TH count was presumed to be under 10,000, sample size correction was applied and we adjusted for potential non-responses by assuming a 10% non-response, leading to our final sample size of 192^23^. Our study encompassed the Fentale district, which consists of 18 sub-districts, excluding the Metehara city. The inclusion of the sedentary villages in the sub-districts was crucial, as the THs there provided healing service to both the pastoralist and settled communities.

The following formula was used to calculate the sample size:

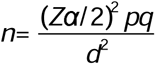

where:

n= minimum sample size

Zα/2 = 1.96 (95% CI) =

P=74.3%, taken from a community based study on knowledge and perception of pastoralist community in the middle and lower Afar area, Ethiopia

q = 1-P = 0.257

d= 5% margin of error

Therefore, the value of n was calculated to be ∼293.

As the number of the study population was estimated to be less than 10,000 in the district, the sample size was corrected and considering a 10% non-response rate, the final sample size was estimated to be approximately 192 THs.

### Patient and Public Involvement statement

Patients and/or the public were not involved in the design, conduct, reporting, or dissemination plans of our research. However, the views and insights of traditional healers, who are key community health stakeholders in the Kereyu pastoralist area, were explored.

### Data Collection Process

Interviews were conducted in person using semi-structured questionnaires. These instruments drew inspiration from past research^6, 24, 25^, but were adapted to suit the local context. Initially crafted in English, the questionnaire was translated into Afan-Oromo (the region’s language) and later back-translated into English to ensure consistency. The questionnaire captured various details: sociodemographic data, knowledge about TB (its causes, symptoms, transmission, prevention, severity, treatment), attitude, practice, and information sources related to TB. Seven proficient data collectors and three regional coordinators who were fluent in the local language undertook the data collection and its oversight. The principal investigator coordinated the field work. Prior to actual data collection, we tested the questionnaire to ensure its clarity, understandability, and reliability. More details on the tools are provided in Supplementary File 1.

### Knowledge Assessment of THs on TB

We evaluated the THs’ knowledge about TB through 19 questions (available in Supplementary file 1). Correct answers were awarded one point, indicating positive knowledge, while incorrect answers were scored zero, indicating a gap in knowledge. The cumulative score ranged between 0 and 19. We used the median score as a threshold, dividing the participants into those with high or low overall TB knowledge. Participants who scored 13 or above (out of a possible 18) were marked as having high knowledge, while those below this threshold were categorized as having low knowledge. For further analysis, only variables showing significant association (p<0.05) in univariate analysis were incorporated into multivariable logistic regression analysis.

### THs’ Attitude Towards TB

We evaluated the THs’ attitude towards TB through seven questions. Responses were categorized as “yes” or “no” for analysis. A “yes” signified a correct attitude towards TB, earning one point, while a “no” suggested a discrepancy in attitude, receiving a zero. We then detailed the proportion of each answer.

### Diagnosis and Treatment Practices Assessment

THs’ current practices regarding TB diagnosis and treatment were assessed via 13 questions. Each answer was converted to either “yes” or “no”, with the percentage of each response type being discussed and described.

### Perception on Collaboration with Formal Health Systems

To understand how THs perceive collaborating with the standard health system, we employed five specific questions. Answers were labeled either “yes” or “no”, with the frequency of each kind of response being elaborated upon and examined.

### Data Analysis

We used the statistical software SPSS (version 22) and STATA to enter and analyze the data. Descriptive statistics helped us summarize the social and demographic status of traditional healers (THs), their knowledge and attitudes towards tuberculosis (TB), and their current practices in diagnosing and treating TB. We reported the number and percentage of respondents in each section. The results are shown in tables and figures.

To find any association between TB knowledge and social and demographic variables, we used the chi-square test, univariate, and multivariable logistic regression analyses. We presented the strength of this association with a 95% confidence interval and a p-value (p<0.05) for the odds ratio.

## Results

### Sociodemographic characteristics of the respondent

From the Fentale district, 268 traditional healers (THs) participated in the study, with 80.6% male and 19.4% female, achieving a 100% response rate. The largest group (28.7%) of participants was aged between 35-44 years. In terms of occupation, most of the THs were agro-pastoralists (61.2%), followed by pastoralists (35.15%). Regarding their education, a large majority (82.5%) could not read and write, while 9.0% had basic literacy skills. A significant number of THs (88.8%) migrated during the dry season, with 86.6% staying within the district. Detailed socio-demographic data is shown in Table 1 below.

**Table 1:** Assessments of the sociodemographic characteristics of the THs of the Kereyu pastoralists, Ethiopia.

| Variables | Percentage % |
| --- | --- |
| **Age** |  |
| 18-24 | 11.6 |
| 25-34 | 28.0 |
| 35-44 | 28.7 |
| 45-54 | 20.5 |
| >55 | 11.2 |
| **Gender** |  |
| Male | 80.6 |
| Female | 19.4 |
| **Marital status** |  |
| Single | 8.6 |
| Married | 88.8 |
| Others (widowed, divorced) | 2.6 |
| **Ethnicity** |  |
| Oromo | 98.5 |
| Amhara | 1.5 |
| **Educational status** |  |
| Unable to read and write | 82.5 |
| Read and write | 9.0 |
| Primary and above | 8.5 |
| **THs category** |  |
| Herbalists | 47.8 |
| Religious or spiritual | 18.7 |
| Casting spells | 17.8 |
| TBA | 8.2 |
| Bonesetters | 7.5 |
| **Religion** |  |
| Muslim | 94.0 |
| Others (Christian, waqefeta) | 6 |
| **Occupation** |  |
| pastoralist | 35.1 |
| agro-pastoralist | 61.2 |
| Others (small business, farming, housewives) | 3.7 |
| **Do you migrate seasonally** |  |
| Yes | 88.8 |
| Within the district | 86.6 |
| Out of the district | 2.2 |

### THs’ Knowledge about TB

The study results show that traditional healers (THs) among the Kereyu pastoralists are familiar with tuberculosis (TB). A vast majority (97.4%) of the THs surveyed acknowledged having heard of TB. Their primary sources of TB information were family members (47.3%), followed by healthcare providers (35.0%). A small percentage mentioned other sources such as meetings and reading materials (2.2%).

Regarding the causes of TB, over half of the THs (52.6%) identified bacteria/germs as the cause of TB. Other significant factors mentioned include close contact with active TB patients (80.2%), living in poorly ventilated houses (58.6%), being HIV positive (37.7%), and experiencing food shortages (29.9%).

An interesting observation was made concerning the differences in knowledge about the causes of TB between THs from sedentary villages (Alga and G/Dima) and those from the pastoralist village (D/Hadu). A significantly higher proportion of THs from the sedentary villages attributed TB to bacteria/germs compared to those from D/Hadu (64.4% vs. 13.0%, p<0.001). There were no significant differences in knowledge observed among other villages.

In terms of symptoms, the majority of THs recognized a persistent cough lasting two weeks or more (87.3%), coughing up blood (80.2%), and weight loss (73.9%) as the primary indicators of TB. Additionally, 96.6% of the THs understood TB to be contagious, with 33.6% specifically mentioning droplets from coughing or sneezing by an active TB patient as the transmission method.

All THs acknowledged the curability of TB, with 73.9% citing medication from healthcare facilities as a treatment option. Interestingly, 30.6% also considered traditional medicine as a viable treatment for TB.

The results from the logistic regression analysis, shown in Table 3, looked at different factors to see how they relate to the traditional healers’ (THs) knowledge about TB. We found that where the THs live, their job, whether they move around with the seasons, and how they learned about traditional medicine (TM) significantly affect their understanding of TB.

**Table 2:** Assessments of knowledge towards TB among the THs of Kereyu pastoralists, Ethiopia.

| **Variables** | **Percentage %**<br>**(n=268)** |
| --- | --- |
| **Heard about TB** | 97.4 |
| **Symptoms of TB** |  |
| Cough lasting > 2 weeks | 87.3 |
| Coughing blood | 80.2 |
| Weight loss | 73.9 |
| Chest pain | 13.4 |
| Weakness | 36.2 |
| Shortness of breath | 68.3 |
| Fever/Sweating | 33.6 |
| **Others (rash, do not know)** | 1.5 |
| **Mode of transmission** |  |
| Droplet through coughing/sneezing | 33.6 |
| Handshaking | 4.1 |
| Sharing utensils | 57.8 |
| Sharing cups | 75.0 |
| Drinking raw milk | 60.8 |
| Eating raw meat | 66.4 |
| Kissing | 42.5 |
| Others (exposure to wind, sharing clothes) | 2.2 |
| **Who can be infected with TB** |  |
| **Anybody** | 89.2 |
| **Only an alcoholic** | 10.4 |
| Others (smokers, khat chewing, HIV positive, etc. ) | 5.2 |
| **TB prevention** |  |
| Covering mouth and nose while coughing /sneezing | 81.0 |
| Using a separate room | 61.9 |
| Avoiding raw milk/meat | 51.9 |
| Avoiding sharing utensils | 54.9 |
| Good nutrition | 46.3 |
| Prayer | 32.5 |
| Others (TB treatment, avoid kissing, good clothing) | 19.4 |
| **TB is curable** |  |
| **Yes** | 99.6 |
| **No** | 0.4 |
| **TB is treated by** |  |
| Drugs provided by healthcare facilities | 73.9 |
| Traditional medicine | 30.6 |
| I don't know | 1.1 |
| **TB drugs are available for free** |  |
| **Yes** | 68.3 |
| **Who can transmit TB** |  |
| Active TB patients | 97.0 |
| Anyone who has a cough | 18.7 |
| Others (undiagnosed TB patients, previously treated) | 2.6 |

\*Note: The sum of % in each category is over 100% due to multiple response options in the questionnaire.

**Table 3:**
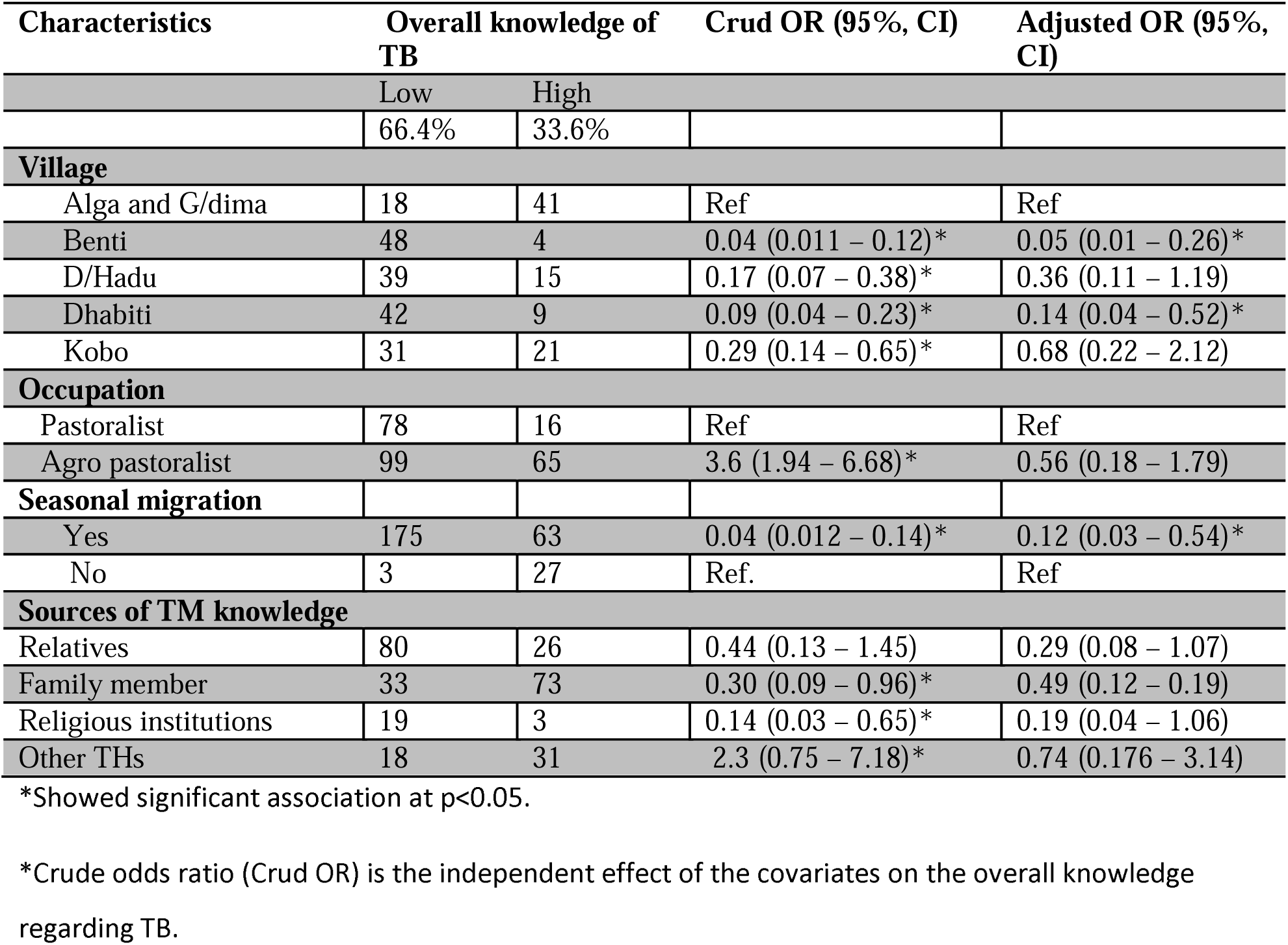
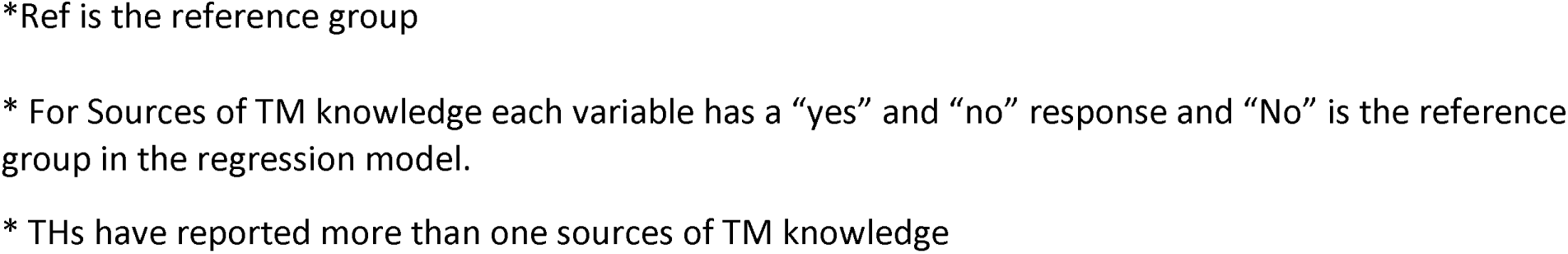
Assessments of THs’ sociodemographic associated with overall knowledge regarding TB in the Kereyu pastoralists’ district in Oromia, Ethiopia.

| Characteristics | Overall knowledge of TB |  | Crud OR (95%, CI) | Adjusted OR (95%, CI) |
| --- | --- | --- | --- | --- |
|  | Low | High |  |  |
|  | 66.4% | 33.6% |  |  |
| **Village** |  |  |  |  |
| Alga and G/dima | 18 | 41 | Ref | Ref |
| Benti | 48 | 4 | 0.04 (0.011 – 0.12)* | 0.05 (0.01 – 0.26)* |
| D/Hadu | 39 | 15 | 0.17 (0.07 – 0.38)* | 0.36 (0.11 – 1.19) |
| Dhabiti | 42 | 9 | 0.09 (0.04 – 0.23)* | 0.14 (0.04 – 0.52)* |
| Kobo | 31 | 21 | 0.29 (0.14 – 0.65)* | 0.68 (0.22 – 2.12) |
| **Occupation** |  |  |  |  |
| Pastoralist | 78 | 16 | Ref | Ref |
| Agro pastoralist | 99 | 65 | 3.6 (1.94 – 6.68)* | 0.56 (0.18 – 1.79) |
| **Seasonal migration** |  |  |  |  |
| Yes | 175 | 63 | 0.04 (0.012 – 0.14)* | 0.12 (0.03 – 0.54)* |
| No | 3 | 27 | Ref. | Ref |
| **Sources of TM knowledge** |  |  |  |  |
| Relatives | 80 | 26 | 0.44 (0.13 – 1.45) | 0.29 (0.08 – 1.07) |
| Family member | 33 | 73 | 0.30 (0.09 – 0.96)* | 0.49 (0.12 – 0.19) |
| Religious institutions | 19 | 3 | 0.14 (0.03 – 0.65)* | 0.19 (0.04 – 1.06) |
| Other THs | 18 | 31 | 2.3 (0.75 – 7.18)* | 0.74 (0.176 – 3.14) |

\*Showed significant association at p<0.05.
\*Crude odds ratio (Crud OR) is the independent effect of the covariates on the overall knowledge regarding TB.
\*Ref is the reference group
\* For Sources of TM knowledge each variable has a “yes” and “no” response and “No” is the reference group in the regression model.
\* THs have reported more than one sources of TM knowledge

Being an agro-pastoralist (someone who farms and keeps livestock) is linked to better knowledge about TB compared to being a nomadic pastoralist (someone who moves livestock from place to place). Specifically, agro-pastoralists were about 3.6 times more likely to have good TB knowledge (COR=3.6, 95% CI= 1.94 – 6.68, P<0.001).

Our analysis also showed that THs who move seasonally tend to know 88% less about TB compared to those who don’t move, even after we consider their job, how they learned about TM, and where they live (adjusted OR=0.12, 95% CI= 0.03-0.05, P=0.021).

In summary, where THs live, their job, their migration habits, and how they got their traditional medical knowledge all play a role in their overall understanding of TB. This is detailed with the odds ratios for each factor in Table 3.

### The THs’ Attitude Towards TB

Most of the traditional healers (THs) surveyed, 84.3%, consider TB to be a very serious disease overall, with 72.0% viewing TB as a particularly serious issue in their own communities. About 38.4% of the THs reported feelings of sadness and hopelessness at the thought of having TB themselves (Table 4). A significant majority, 85.5%, believe that individuals who are HIV-positive should be particularly concerned about TB. Additionally, nearly half of the THs, 48.5%, feel that TB is viewed with less stigma than HIV/AIDS within their community.

**Table 4:**
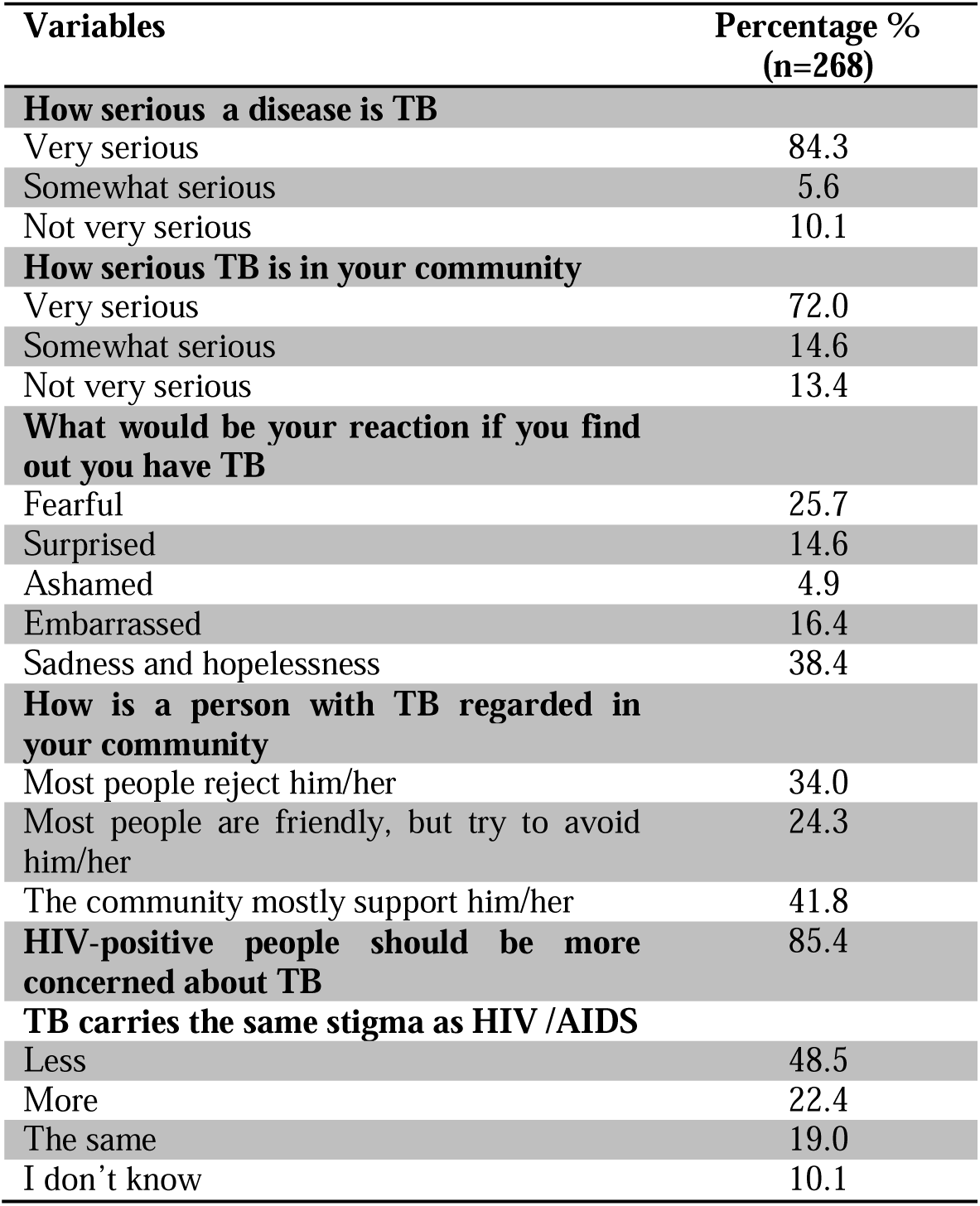
Assessments of attitude towards TB among the THs of Kereyu pastoralists in Ethiopia.

| Variables | Percentage %<br>(n=268) |
| --- | --- |
| **How serious a disease is TB** |  |
| Very serious | 84.3 |
| Somewhat serious | 5.6 |
| Not very serious | 10.1 |
| **How serious TB is in your community** |  |
| Very serious | 72.0 |
| Somewhat serious | 14.6 |
| Not very serious | 13.4 |
| **What would be your reaction if you find out you have TB** |  |
| Fearful | 25.7 |
| Surprised | 14.6 |
| Ashamed | 4.9 |
| Embarrassed | 16.4 |
| Sadness and hopelessness | 38.4 |
| **How is a person with TB regarded in your community** |  |
| Most people reject him/her | 34.0 |
| Most people are friendly, but try to avoid him/her | 24.3 |
| The community mostly support him/her | 41.8 |
| **HIV-positive people should be more concerned about TB** | 85.4 |
| **TB carries the same stigma as HIV /AIDS** |  |
| Less | 48.5 |
| More | 22.4 |
| The same | 19.0 |
| I don't know | 10.1 |

### TB Diagnosis and Treatment Practices Among THs

Most traditional healers (THs) (91.45%) believe that the community trusts traditional medicine (TM) because it is effective (60.4%), easily accessible (45.5%), and affordable (29.9%). The symptoms they most commonly used to determine if someone has TB include a cough that lasts for three weeks (71.5%), coughing up blood (68.0%), and weight loss (59.5%). A significant number (66%) of THs reported referring someone showing symptoms of TB to a medical facility (Table 5).

**Table 5:** Assessments of TB diagnosis and treatment practice of the THs of Kereyu pastoralists, Ethiopia.

| Variables | Percentage % (n=268) |
| --- | --- |
| **The number of TB cases increasing** |  |
| Yes | 41.8 |
| No | 58.2 |
| **How do you diagnose TB** |  |
| Symptoms | 39.9 |
| Results from a healthcare facility | 28.7 |
| Physical examination | 9.0 |
| Symptoms and physical examination | 41.8 |
| Previous history | 1.9 |
| **Physical examinations** |  |
| Fever | 13.4 |
| Skin color change | 78.7 |
| Others (swelling, rash, discharge, etc.) | 19.4 |
| **How do you treat a person with TB** |  |
| Casting spells | 17.9 |
| Treat with plants/herbal medicine | 47.8 |
| Prayer and holy water | 18.7 |
| Refer to a healthcare facility | 7.1 |
| Others (home rest, honey with milk, recommend good nutrition, etc.) | 8.6 |
| **How often a patient gets cured of TB with your medication** |  |
| Always | 9.3 |
| Most of the time | 54.9 |
| Sometimes | 27.6 |
| Rarely | 3.0 |
| Never | 4.1 |
| Others (I don't know) | 1.1 |
| **The advice given to a patient** |  |
| Transmission prevention | 74.6 |
| Balanced diet | 39.6 |
| No advice | 3.4 |
| Others (follow treatment properly, home rest, no mixing up treatment, etc.) | 19.4 |
| **Ever referred TB patient to a health facility** |  |
| Yes | 66.0 |

### Perceptions of THs on Collaboration with the Conventional Health System

More than half of the traditional healers (THs) (54.1%) favor the conventional health system, with 29.5% preferring a combination of both modern and traditional medicine (TM) for healthcare services, while 16.4% prefer TM alone. A vast majority (86.2%) of THs are open to collaborating with the conventional health system. Additionally, 85.15% support the idea of integrating traditional and modern medicine. Suggested areas for collaboration include working together on diagnosis and treatment (61.2%), cross-visiting between practitioners (60.4%), and learning about modern medicine (57.7%) (Table 6).

**Table 6:**
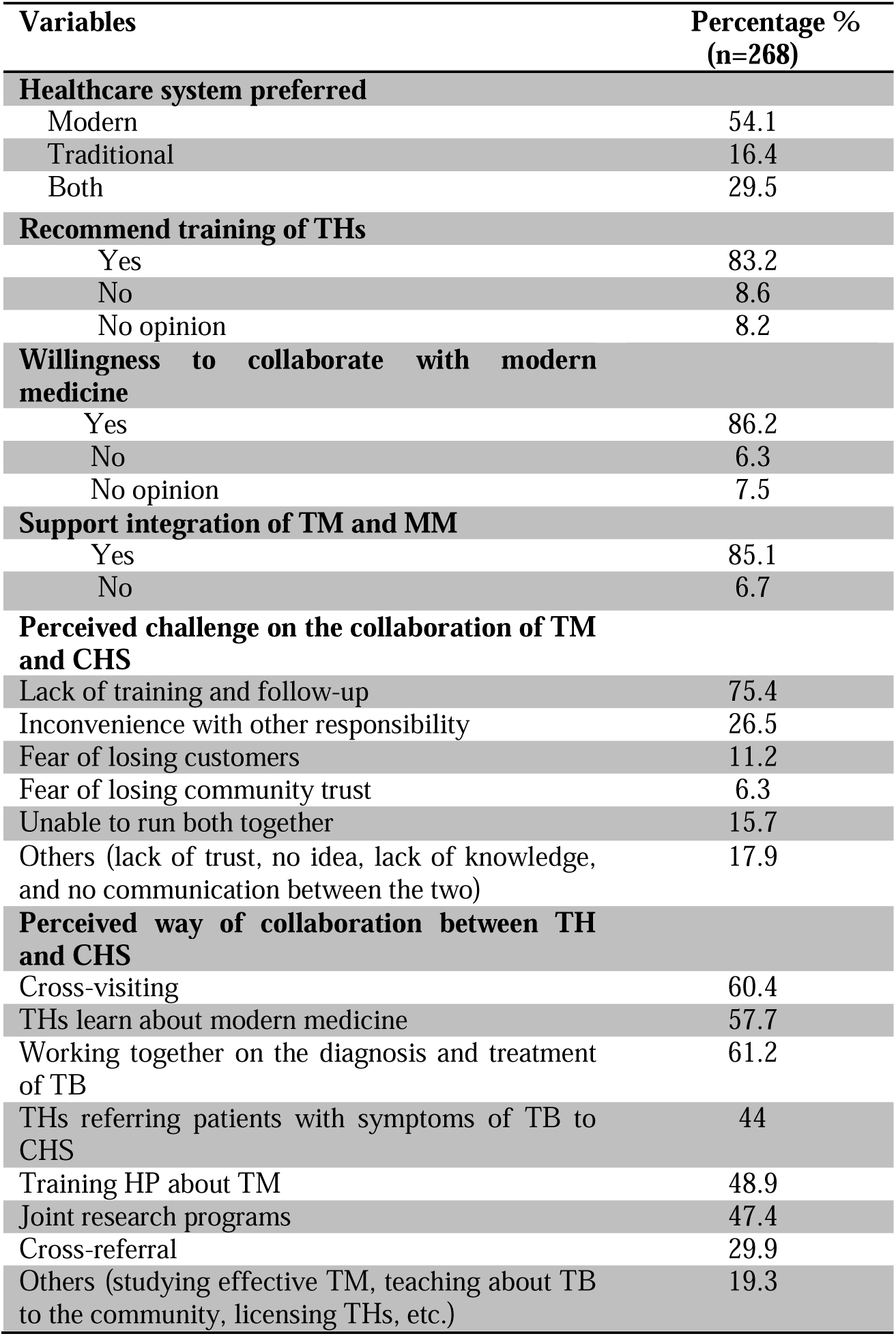
Assessments of the THs’ perception about collaboration with the conventional health system, Kereyu, Ethiopia.

| Variables | Percentage %<br>(n=268) |
| --- | --- |
| **Healthcare system preferred** |  |
| Modern | 54.1 |
| Traditional | 16.4 |
| Both | 29.5 |
| **Recommend training of THs** |  |
| Yes | 83.2 |
| No | 8.6 |
| No opinion | 8.2 |
| **Willingness to collaborate with modern medicine** |  |
| Yes | 86.2 |
| No | 6.3 |
| No opinion | 7.5 |
| **Support integration of TM and MM** |  |
| Yes | 85.1 |
| No | 6.7 |
| **Perceived challenge on the collaboration of TM and CHS** |  |
| Lack of training and follow-up | 75.4 |
| Inconvenience with other responsibility | 26.5 |
| Fear of losing customers | 11.2 |
| Fear of losing community trust | 6.3 |
| Unable to run both together | 15.7 |
| Others (lack of trust, no idea, lack of knowledge, and no communication between the two) | 17.9 |
| **Perceived way of collaboration between TH and CHS** |  |
| Cross-visiting | 60.4 |
| THs learn about modern medicine | 57.7 |
| Working together on the diagnosis and treatment of TB | 61.2 |
| THs referring patients with symptoms of TB to CHS | 44 |
| Training HP about TM | 48.9 |
| Joint research programs | 47.4 |
| Cross-referral | 29.9 |
| Others (studying effective TM, teaching about TB to the community, licensing THs, etc.) | 19.3 |

## Discussion

This study revealed that most traditional healers (THs) are aware of tuberculosis (TB) and have a basic understanding of its symptoms. However, a significant knowledge gap exists regarding the biomedical causes of TB. The study also identified misconceptions about how TB is transmitted, factors that increase one’s risk of contracting TB, and the use of traditional medicine (TM) to treat TB. Notably, a significant number of THs expressed a willingness to work together with the conventional healthcare system to control TB.

Traditional beliefs, such as hard work, cold air, malnutrition, sexual overindulgence, dust, and witchcraft, were frequently cited as causes of TB in this study. Similar perceptions were also noted among the Somali, Afar and Shinille pastoralists in Ethiopia^22, 23^, as well as in other community-based studies in the study area^19^ and within the country^26^. This limited understanding of the causes of TB may result in traditional healers (THs) turning to traditional medicine (TM) for treatment. While this approach may not effectively cure TB, it could contribute to the further spread of the disease within the community.

In this study, traditional healers (THs) identified sharing cups and utensils, consuming raw milk and meat, and kissing as primary routes of TB transmission, aligning with findings from studies in Afar and Shinille pastoralists in Ethiopia^5, 22–24^ a,mjkløænd among pastoralist community in Kenya7. Such misconceptions about the route of TB transmission and factors exposing people to the disease could lead to a negative attitude towards TB patients and may also result in poor preventive and control efforts. However, most THs also practice preventative measures like covering their mouth when coughing and not sharing utensils, showing better awareness than seen in some past studies^23–25, 27^.

Our analysis found that THs with stable living conditions, agro-pastoral occupations, less seasonal migration, and familial knowledge sources had a better understanding of TB. This suggests that the nomadic lifestyle may limit access to accurate health information, affecting TB control efforts ^17, 20^. Early consultation with THs by TB patients has been linked to delays in diagnosis and treatment^6, 11, 20^, underscoring the need for better integration between traditional and formal healthcare practices.

Regarding attitudes, the seriousness of TB is well recognized among THs, likely due to the volume of patients with chronic TB symptoms seeking their services. This recognition, however, may also lead to stigmatization within the community, especially for those with chronic symptoms.

Notably, a significant portion of THs consider traditional medicine a viable TB treatment, often using herbs and plants. This perception, higher than in other studies^11^, could discourage proper healthcare seeking, potentially worsening TB outcomes and facilitating the spread of the disease, including drug-resistant strains^28, 29^.

Despite these challenges, a strong willingness exists among THs to collaborate with the conventional healthcare system on TB control, a sentiment echoed by healthcare providers in the study area ^30^ and similar studies in other settings^13, 14, 16, 29, 31^. This mutual interest presents an opportunity for the national TB control program to foster collaboration, although challenges like mistrust and lack of consensus remain.

Limitations include potential bias in self-reported practices by THs and the absence of a baseline survey due to financial constraints. However, the high response rate and comprehensive inclusion of THs across the district strengthen the study’s reliability. This pioneering study in Ethiopia highlights the potential for integrating THs into conventional TB control efforts within pastoralist communities. While we are acknowledging the time gap between the data collection in 2015 and the current submission for publication, it is important to underscore the potential implications this may have on the validity of our findings. However, we have determined that the data remains highly relevant for several reasons. Firstly, no substantial changes have occurred in the study area concerning the epidemiology, treatment, or control strategies for tuberculosis that would undermine the applicability of our results to the current context. This suggests that the insights offered by our research are likely to retain their value over time. Furthermore, the aspects of human behavior, cultural practices, and perceptions explored in our study are typically subject to slower change and have not been significantly influenced by major societal shifts since the data collection. Lastly, there is an evident scarcity of recent studies in the area, implying that our data has the potential to bridge a critical gap in the literature. This study, therefore, contributes valuable insights that have not been overtaken by new research, reinforcing the pertinence and contribution of this study to the field.

## Conclusions and Recommendations

This study identified significant knowledge gaps among THs regarding the causes and transmission of TB. Misconceptions about TB can negatively impact TB control efforts. However, the willingness of THs to collaborate with the healthcare system presents a promising opportunity. Future efforts should focus on educating THs about TB and establishing effective collaboration strategies to improve TB control in pastoralist communities.

## Data Availability

All data produced in the present work are contained in the manuscript.

## Lists of abbreviations

DOTS: Directly observed treatment short course
FGD: Focus group discussion
FMoH: Federal Minster of Health
HCP: Healthcare providers
HEWs: Health Extension Workers
HEP: Health extension program
HIV: Human immunodeficiency virus
HIV/AIDS: Human immunodeficiency virus/ acquired immunodeficiency syndrome
TB: Tuberculosis
THs: Traditional healers
TM: Traditional Medicine
NSD: The Norwegian Social Science Data Service
SPSS: Statistical Software for Social Science
WHO: World Health Organization

## Declarations

### Ethical approval and consent to participate

The Norwegian Social Science Data Service (NSD) and Ethical Review Committee of Jimma University, Jimma Ethiopia and the Oromia Regional Health Office Ethical Review Committee, Addis Ababa, Ethiopia approved this study. The THs provided written consent to participate after receiving information about the study. We used codes instead of personal identifiers to maintain the confidentiality and anonymity of the study participants.

### Consent to publish

Not applicable

### Competing interest

The authors declare no competing interests.

### Availability of data and material

All data sets generated or analyzed during the current study were included in this manuscript.

## Funding

This was a PhD project and was supported by the single student support program at the University of Oslo.

## Authors’ contributions

BTS developed the study, prepared the questionnaire, collected, analyzed and interpreted the data, and wrote the manuscript. FA, TB, and GB critically revised the manuscript. All the authors have read and approved the final manuscript.

## Acknowledgements

The study was financially supported by the University of Oslo. We would like to thank the Oromia Regional Health Office for permission to conduct the study and the Fentalle District Health Office for facilitating the fieldwork. Last but not least, we would like to thank our respondents for their participation in the study.

## Questionnaire

Supplementary File 1

## Notes

### Competing Interest Statement

The authors have declared no competing interest.

### Author Declarations

1.The Norwegian Social Science Data Service (NSD) and 2. Ethical Review Committee of Jimma University, Jimma Ethiopia and 3. The Oromia Regional Health Office Ethical Review Committee, Addis Ababa, Ethiopia approved this study.

